# Effect of introducing interprofessional education concepts on students of various healthcare disciplines in the United Arab Emirates

**DOI:** 10.1101/2021.04.02.21254859

**Authors:** Shroque Zaher, Farah Otaki, Nabil Zary, Amina Al Marzouqi, Rajan Radhakrishnan

## Abstract

**Background:** The value of interprofessional education (IPE) in nurturing healthcare professionals and shaping their professional identities and attitudes towards interdisciplinary teamwork and collaboration is established in the literature. It is an emerging concept in the Middle East and North Africa region and is new to the United Arab Emirates (UAE).

**Purpose:** To investigate the effect of one of the first experiences of IPE in the UAE, which was designed in alignment with the principles of the Situated Learning Theory, on students of various healthcare disciplines readiness for interprofessional learning and collaboration.

**Methods:** A pre-post intervention quantitative research design was adopted for this study to assess the effectiveness of the intervention in raising the level of readiness for interprofessional work among the participants. The participants included students of medicine, pharmacy, nursing, and physiotherapy. Readiness for Interprofessional Learning Scale (RIPLS) was used as the pre- and post-intervention assessments; data was analysed using SPSS.

**Results:** The IPE intervention under investigation significantly increased the level of readiness to engage in cross-disciplinary learning and collaboration among participating health professions’ students. In terms of the subscales, the participants’ openness to engage in teamwork was raised and their professional identity was fostered. Yet, no statistical significance around clarity of roles and responsibilities was detected.

**Conclusion:** IPE interventions hold the potential to significantly increase receptiveness to cross-disciplinary learning and collaboration among health professions’ students. The findings of this study encourage other universities in the MENA region to adopt IPE to improve future health professionals’ capacity to develop shared understanding and mutual respect within cross-disciplinary teams, which ultimately feed into improved quality of care and patient outcome.

## Introduction

Interprofessional education (IPE) is an inter-collaborative approach to develop healthcare students as future interprofessional team members, and is a practice promoted by the World Health Organisation (WHO) as well as by various international organisations such as the Canadian Interprofessional Health Collaborative (CIHC), the European Interprofessional Education Network, and the UK Centre for the Advancement of Interprofessional Education (1, 2).

Relational Coordination (RC) is a concept that appears to be central to IPE. It is defined as a mutually-reinforcing process of interaction between communication and relationships carried out for the purpose of task integration (3). The RC theory suggests that for a team to be effectively coordinated, there is a need for the development of shared knowledge and understanding among its team members, as well as for the team’s relationships to be built on shared goals and mutual respect (4). This highlights the importance of communication and relating to one another for the purpose of task integration (5). Relationships help shape the communication through which coordination occurs, and hence it is of utmost importance that emphasis is placed on teamwork and collaboration in the training of students in healthcare professions.

Today’s patients have complex health needs and typically require more than one discipline to address issues regarding their health status to attain improved patient outcomes. Many of the patients’ adverse events and poor health outcomes are attributed to misunderstanding or poor communication among members of the interprofessional teams. Even though physicians and nurses work together, their academic courses are separate and the training in effective strategies of communication and care is often postponed to future professional practice (1, 6). Studies support the introduction of interprofessional education at the start of healthcare student’s professional education rather than at the end (7, 8). This also applies to attempts to prevent the formation of negative interprofessional attitudes.

Another theory that is central to our study is the Situated Learning Theory (SLT) which was initially proposed by Jean Lave and Etienne Wenger in the late 1980s. The concept of SLT relates to the conceptualization that learning occurs within authentic context, culture, and activity. It promotes the idea that students learn better in collaborative group settings and when the activities are based on real-life experiences. Knowledge needs to be presented in authentic contexts-settings and situations, that would typically require deploying that knowledge. Social interaction and collaboration are essential components of SLT; learners become involved in a “community-of-practice” which embodies certain beliefs and behaviors to be acquired. As the beginners or novices move from the periphery of a community to its center, they become more active and engaged within the culture and eventually assume the role of experts (9).

It is established that IPE has the potential to positively influence attitudes and perceptions towards interprofessional collaboration and increase the effectiveness of clinical decision-making and ultimately the quality of care (10, 11). Many of the key competencies for IPE relate primarily to teamwork, including skills in setting shared goals, communication, knowledge of roles and responsibilities, and negotiation for conflict resolution (1, 5, 12). Understanding one’s own role and that of other professionals in the healthcare team is critical in IPE. As students become more immersed in their own education, they are likely to gain a better and more comprehensive understanding of their role in the interdisciplinary healthcare team. Establishing a strong IPE foundation and opportunities for interprofessional learning within healthcare education is therefore paramount, as has long been emphasized by the WHO (2, 13). Yet, many health professionals have had little or no exposure to IPE activities during their own training, and many clinical sites in which faculty oversee training lack robust or explicit examples of interprofessional team-based care (14).

Interprofessional collaboration is an emerging concept in the Middle East and North Africa (MENA) region and is new to the United Arab Emirates (UAE) (15). In this paper, we describe one of the first structured experiences of IPE in the UAE, conducted at the Mohammed Bin Rashid University of Medicine and Health Sciences (MBRU) in collaboration with the University of Sharjah (UoS). This intervention was based on the principles of the SLT with the understanding that “real” learning can only happen when it is contextual (i.e., simulating an authentic experience that will engage the learners in complex, realistic, and problem-centered activities while recasting the role of the teacher to a facilitator) (16-18).

Given the value of IPE in nurturing healthcare professionals, shaping their professional identities and attitudes toward interdisciplinary teamwork and collaboration, and ultimately in improving outcomes of care (19), the purpose of this study was to investigate the effect of an IPE intervention on student readiness for interprofessional learning and collaboration.

Accordingly, this study’s research questions are as follows:

- How efficacious was this IPE intervention at raising the level of readiness for interprofessional learning and collaboration among the health professions’ student participants?
- How did this IPE intervention affect the students’ knowledge, skills, and attitudes around their teamwork and collaboration, professional identity, and roles and responsibilities?
- How did the effect of this IPE intervention on the level of readiness for interprofessional learning and coordination vary across different professions?

## Materials and methods

### Context of the study

The study was carried out at MBRU in collaboration with the UoS. MBRU is one of 8 medical colleges in the UAE and one of two in Dubai. MBRU was established in 2016, and currently offers a 6-year Bachelor of Medicine, Bachelor of Surgery (MBBS) program as well as postgraduate degrees in dentistry, nursing, and biomedical sciences. The first 3 years of this MBBS program are preclinical and the remaining years are spent in the clinical setting. Being a young university, MBRU’s vision is focused on innovation and research, not least so in the educational setting. In line with this, one of the longitudinal themes of the MBBS program is enhancing multidisciplinary teamwork in students, which formed the basis of our study.

The UoS was established in 1997 and offers 110 programs across multiple academic disciplines. It offers both undergraduate and postgraduate degrees in medicine, nursing, pharmacy, and other allied health sciences such as physiotherapy. Both universities share a vision of graduating future doctors with the necessary interpersonal skills that would enable comprehensive multidisciplinary patient care.

### Description of the IPE session under investigation

As part of the Neurosciences Course which takes place in the second semester of the third year of the MBBS program of MBRU, all enrolled students (i.e., Class of 2022) were required to attend an IPE session which was designed, implemented, and evaluated in alignment with the respective program outcome of promoting collaborative teamwork amongst students. In line with the principles of SLT (17), the session was based on realistic case-based problems with students split into teams, and the session facilitated by instructors, ensuring that their role was not that of a ‘teacher’ but that of a ‘coach’. Emphasis was placed on fostering meaningful interactions between the students within a cross-disciplinary team setting. The students were split into groups of eight. To each group of MBRU MBBS students, student representation was added from the schools of pharmacy, nursing, and physiotherapy from the UoS. The session was facilitated by two faculty members: a professor of pharmacology (RR) and an assistant professor of pathology (SZ). The session comprised of an initial introduction to the learning concepts of inter-professional healthcare teams and collaborative patient-centered care. The preset learning outcomes were as follows:

- Define interprofessional collaborative practice.
- Discuss the benefits of interprofessional collaborative practice including the impact on quality and safety of patient care.
- Describe key elements of effective interprofessional team-based care.
- Discuss factors that may influence interprofessional collaboration.
- Describe the roles, responsibilities, and abilities of various healthcare professions involved in collaborative work, including their training and respective scopes of practice.
- Describe one’s own professional role in relation to collaborating with other healthcare professionals.

The session then revolved around two patient case-based scenarios, created by the abovementioned faculty members, on the topics of stroke and pain. These scenarios were designed, in alignment with SLT, to ensure that they contain sufficient material to generate discussion across all participating disciplines. The students were asked to discuss the cases and then answer a series of questions which included:

1. What is the immediate management of this patient?
2. What are the long-term sequelae of this event?
3. How can this patient be managed in the long-term and which professions will play a role?
4. How can the effect of another similar event be minimized for this patient?
5. What other health and social concerns are of importance to this patient?
6. What is the long-term management plan of this patient?

### Research Design

A pre-post intervention quantitative research design was adopted for this study to assess the effectiveness of the IPE intervention in raising the level of readiness for interprofessional work among the participants. The study was conducted in two rounds.

Ethical approval for the study was granted by the MBRU, Institutional Review Board (Reference # MBRU-IRB-2020-016).

### Data Collection

The data was collected using the Readiness for Interprofessional Learning Scale (RIPLS) questionnaire. RIPLS (20, 21) is a self-reporting tool that assesses perceptions of healthcare students’ knowledge, skills, and attitudes regarding readiness to learn with other healthcare professionals. It is composed of 19 components that are measured using a 5-point Likert-type scale. The 19 components are divided into three validated subscales. The first one is the “Teamwork/ Collaboration” subscale which assesses the extent to which the participant values cooperative learning and respecting students from other healthcare professionals. The second one is the “Professional Identity” subscale which measures the tendency of the participant to value and benefit from collaborative relationships with other healthcare professionals. As for the third one, it is the “Roles and Responsibilities” subscale, and measures the practical application of interprofessional skills with other healthcare professional students. Two out of those three subscales are further divided into two segments. The Teamwork/ Collaboration subscale has the “Need for positive relationships between professor and other healthcare students” and “Acquisition and effectiveness of teamworking skills” segments. As for the Professional Identity subscale, it has “Positive” and “Negative” segments. Scores can be generated for the segments, subscales, and overall rating. Higher scores indicate more readiness to engage in interprofessional learning experiences. It is worth noting that given the inverse nature of the Negative Professional Identity segment of the tool, the Likert-type scale of its components (10 through 12) were coded as such: 1: Strongly Agree, 2: Agree, 3: Neutral, 4: Disagree, and 5: Strongly Disagree.

## Data Analysis

### Quantitative Analyses

The quantitative data was descriptively analysed using IBM SPSS Statistics Version 27. For each of the demographic variable, the number of cases, and frequencies and valid percentages were calculated. For each of the 19 quantitative components of the tool, the mean and standard deviation were calculated. An overall score of readiness was calculated (1 through 19), along with scores for each of the 3 subscales of the tool: Teamwork and collaboration (1 through 9), Professional identity (10 through 16), and Roles and responsibilities (17 through 19). Additional scores were calculated for the two segments of the Teamwork and collaboration subscale: Acquisition and effectiveness of teamworking skills (1 through 6) and Need for positive relationships between professionals and other healthcare students (7 through 9), and for the two segments of the Professional identity subscale: Negative (10 through 12) and Positive (13 through 16).

The validity tests of Cronbach’s Alpha and the Principal Component Analysis (PCA) were performed to ensure the internal consistency and check external variance, respectively, of the adapted tool.

For the inferential analyses, to select the appropriate tests, a test of normality was conducted for the data of each of the 19 components, and for all eight scores (overall, 3 subscales, and 2 segments within each of 2 of the subscales). The data of each of the 19 components, independently, and all the scores turned out to be not normally distributed. Accordingly, Mann-Whitney tests were used to compare the scores, and each component independently, pre- and post-intervention.

The scores, and each component independently, were compared across the demographic variables, as well: Year of Birth (YoB), Gender, and Discipline. For the dichotomous variable: Gender, Mann-Whitney test was used. As for the other two demographic variables, Kruskal-Wallis tests were conducted.

## Results

### Quantitative Analyses

The response rates of the pre- and post-assessment of both rounds were 85.56% and 93.33%, respectively. In total, 161 responses were collected, out of which 88 were from the 1^st^ round and 73 were from the 2^nd^ round (Table 1).

**Table 1.** Response rates pre- and post-intervention in both rounds

| Round | Pre |  |  | Post |  |  | Total |  |  |
| --- | --- | --- | --- | --- | --- | --- | --- | --- | --- |
|  | Number of Responses | Number of Participants | Response Rate | Number of Responses | Number of Participants | Response Rate | Number of Responses | Number of Participants | Response Rate |
| 1st | 43 | 45 | 95.56 | 45 | 45 | 100 | 88 | 90 | 97.78 |
| 2nd | 34 | 45 | 75.56 | 39 | 45 | 86.67 | 73 | 90 | 81.11 |
| Total | 77 | 90 | 85.56 | 84 | 90 | 93.33 | 161 | 180 | 89.44 |

As illustrated in Table 2, the Year of Birth (YoB) of the responders, who reported on this variable, ranged between 1994 and 2000, with the biggest portion of responders born in year 1999 (31.1% in the pre-assessment and 35.1% in the post-assessment). Most of the responders who reported on their Gender were Female (74.7% in the pre-assessment and 71.1% in the post-assessment). In terms of Discipline, most of the responders were in Medicine (44.2% in the pre-assessment and 47.6% in the post-assessment), followed by Pharmacy, Physiology, and Nursing, consecutively.

**Table 2.**
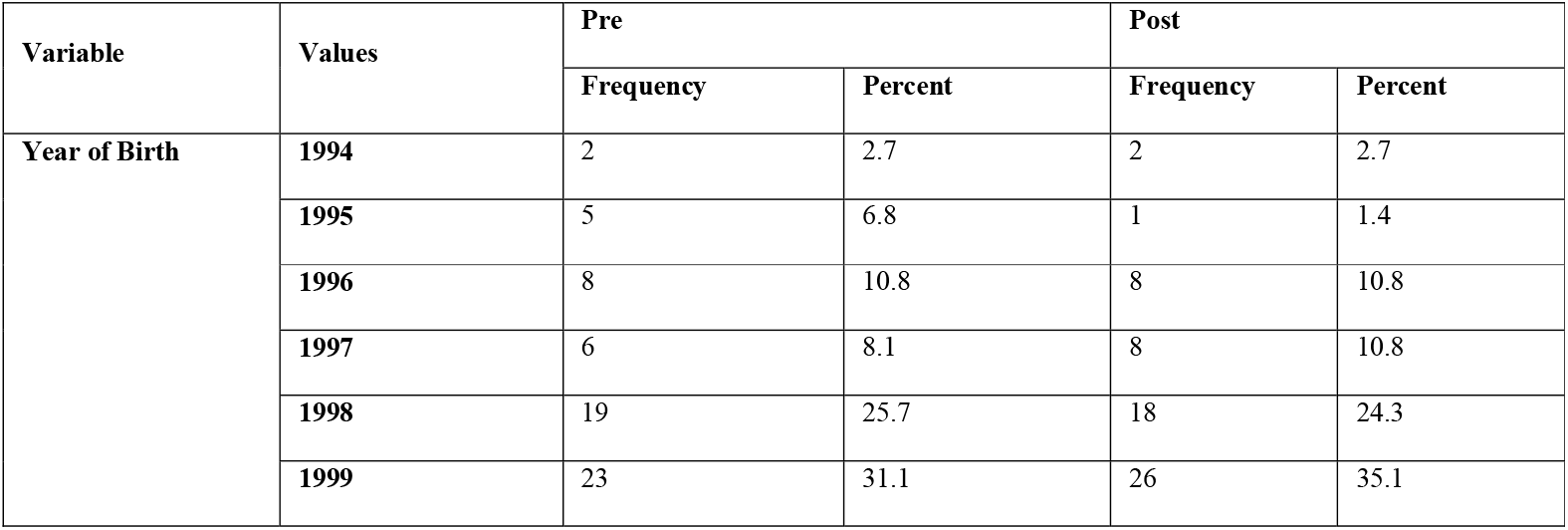
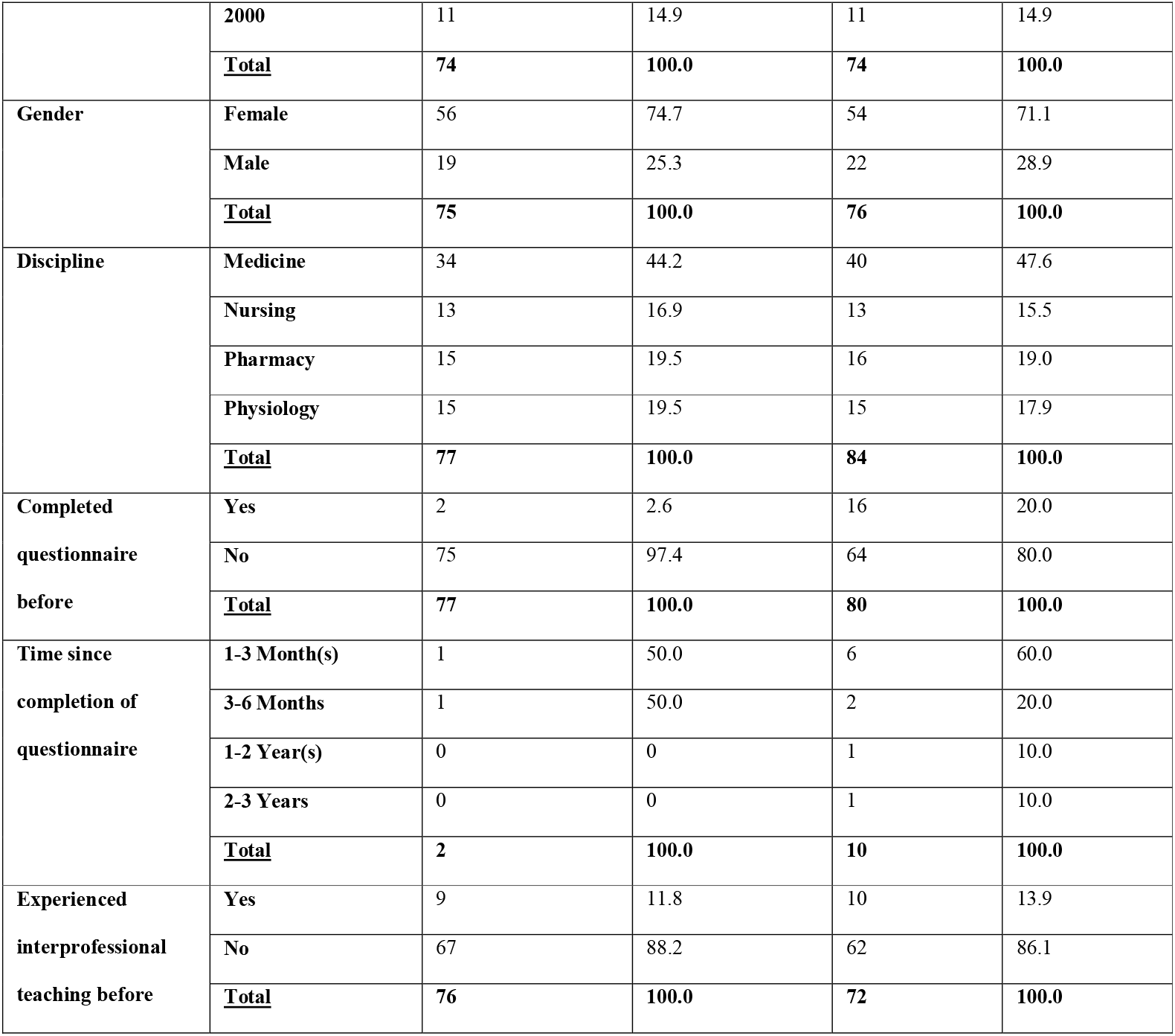
Output of descriptive analysis of demographics data

A total of 75 responders in the pre-assessment and of 64 responders in the post-assessment indicated that this was the first time they complete a RIPLS questionnaire. In the pre-assessment, 1 responder reported completing it 1-3 Month(s) ago and 1 responder reported completing it 3-6 Months ago. As for the post-assessment, 6 responders reported completing it in 1-3 Month(s), 2 in 3-6 Months, and 1 in each of the remaining categories: 1-2 Year(s) and 2-3 Years. For 88.2% of the responders to the pre-assessment and 86.1% of those to the post-assessment, this intervention constituted their first exposure to interprofessional teaching (Table 3).

**Table 3.**
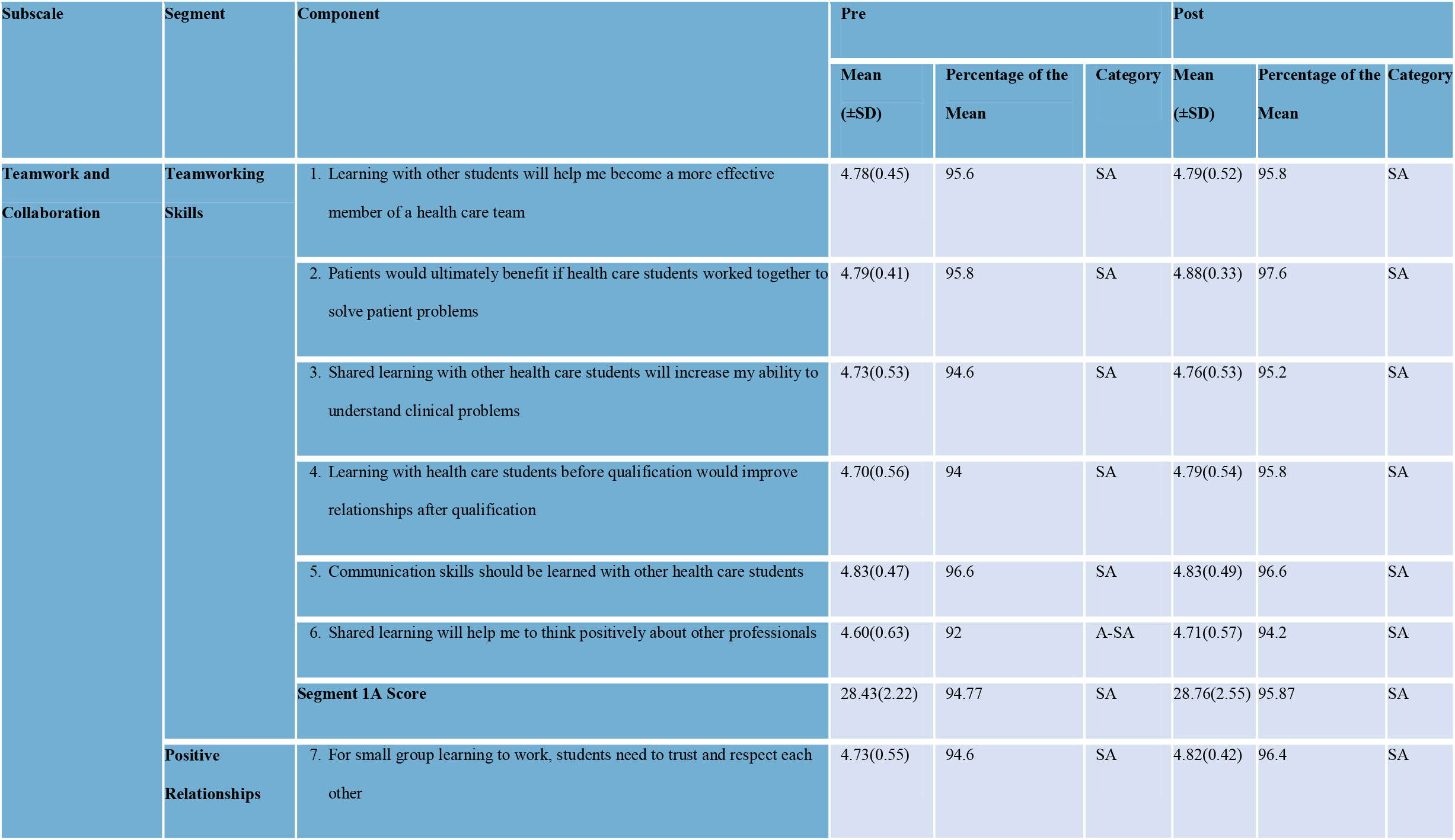
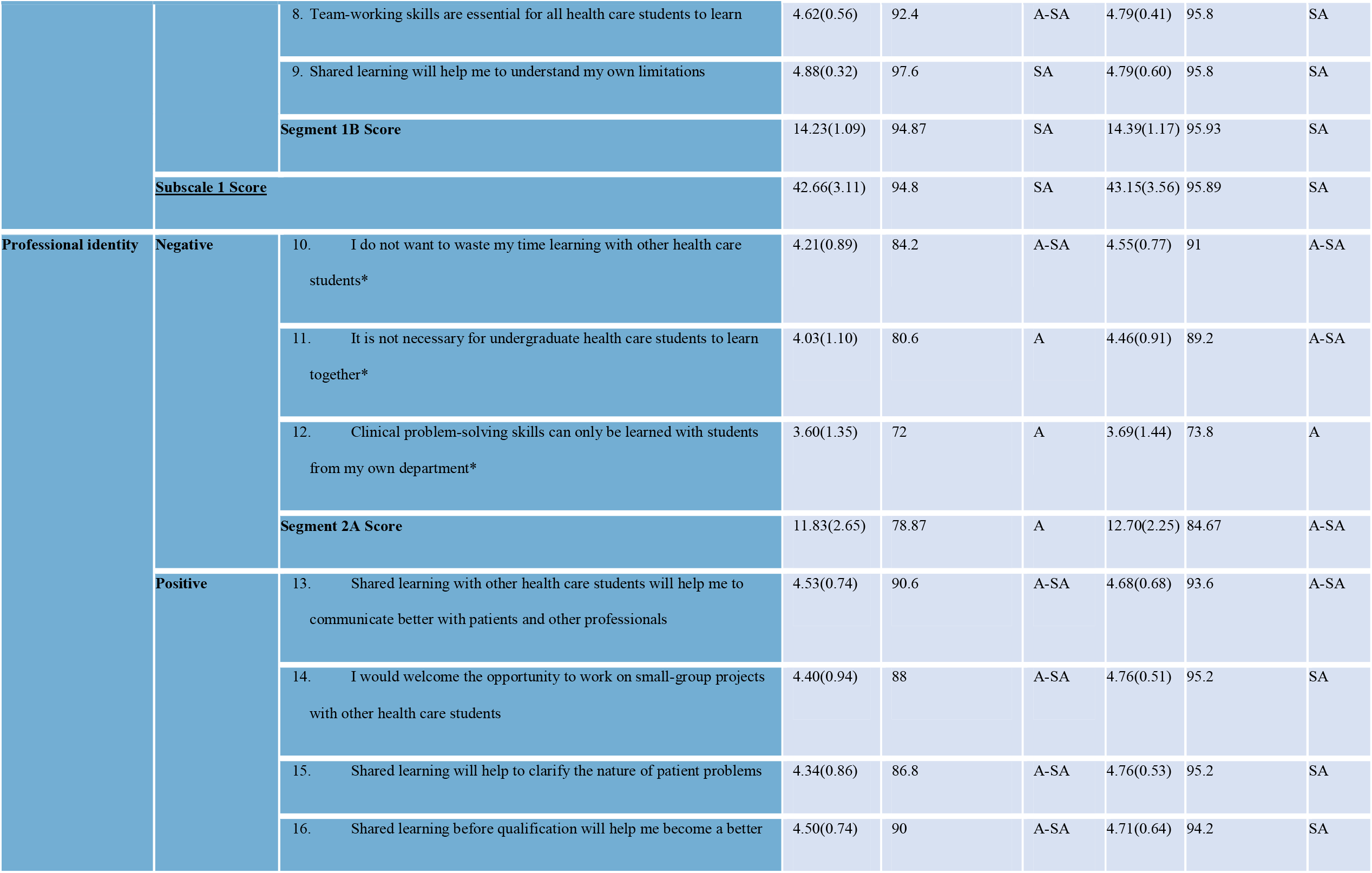
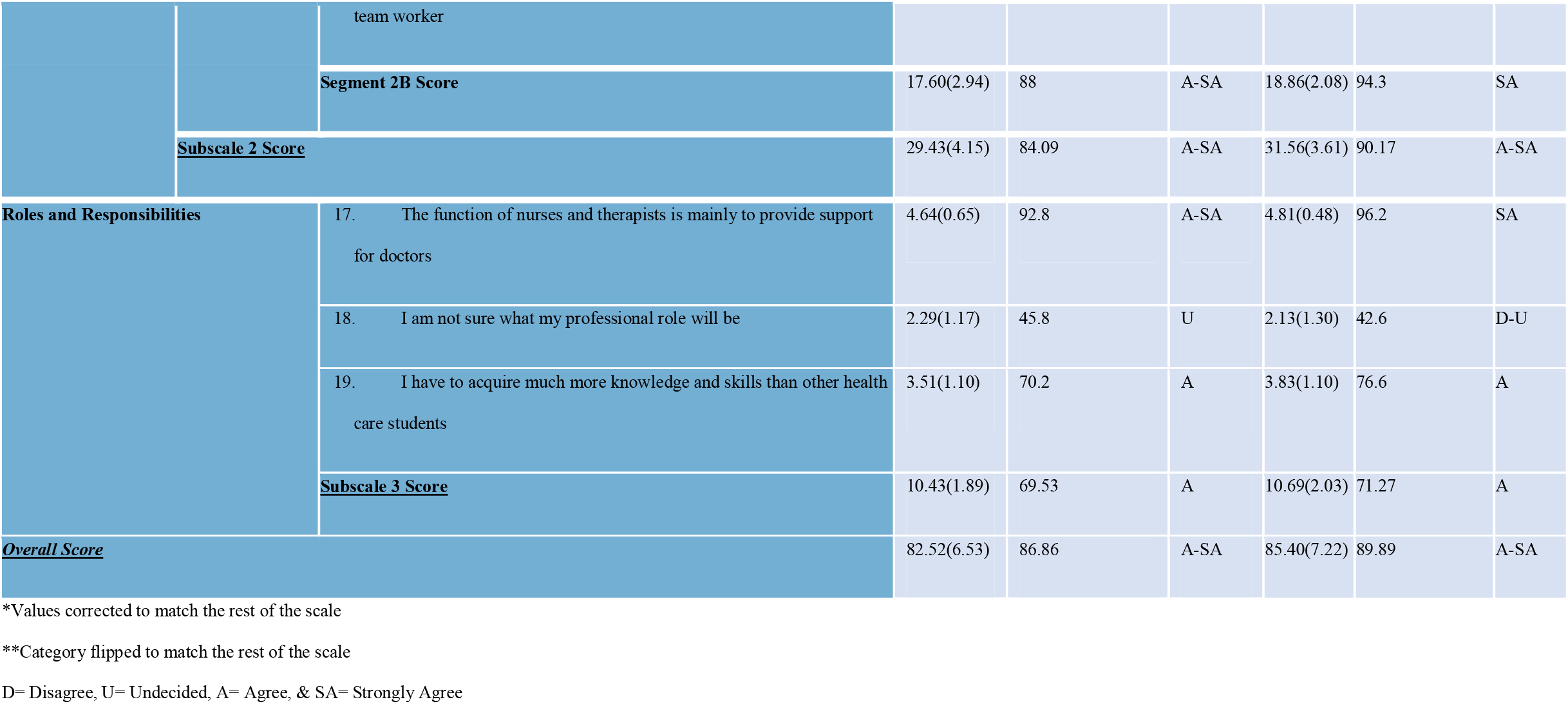
Output of descriptive quantitative analysis of all the components and scores, pre- and post-intervention

The reliability score of Cronbach’s Alpha for the adapted RIPLS questionnaire was 72.4%. The percentage of the overall score was 86.86% pre-intervention and 89.89% post-intervention, as per Table 4. According to the PCA, 87.4% of the variance can be explained by the instrument (P<0.001) which means the instrument is not only reliable but also valid to measure what it is intended to measure.

The post-assessment overall readiness score, with a mean of agreement of 85.40(±7.22), was significantly higher than its pre-assessment counterpart, with a mean of agreement of 82.52(±6.53) (p=0.001).

The Teamwork and collaboration subscale score was significantly higher in the post-assessment, relative to the pre-assessment (p=0.031). The Teamworking skills segment score was also significantly higher in the post-assessment (p=0.037), while there was no significant difference in the Positive relationships segment score. In terms of the components of the Teamwork and collaboration subscale (1 through 9), none showed significant difference between the pre- and post-assessments.

As for the Professional identity subscale score, with a mean of agreement of 31.56(±3.61), it was significantly higher than the same score assessed pre-intervention, with a mean of satisfaction of 29.43(±4.15) (p=0.001). The two segments of this score: Negative and Positive professional identity, were also significantly higher in the post-assessment relative to the pre-assessment (p<0.05). In terms of the components of the Professional identity subscale (10 through 16), components 12 and 13 assessed post-intervention were not significantly different than their pre-intervention assessment. As for the remaining components (10 and 11, and 14, 15, and 16), they significantly increased in the assessment after the intervention (p<0.01).

As for the Roles and responsibilities subscale, the analysis showed no statistically significant difference between pre- and post-assessment; among the components of the respective subscale (17 through 19), only 17 significantly increased in the assessment after the intervention (p=0.034).

In the pre-assessment, the medicine participants scored significantly higher than those of the rest of the disciplines in the Teamworking skills segment. The same category of participants also scored higher, relative to others, in relation to the following components: 5 and 6, and 13 (p<0.05). As for component 2, pharmacy participants scored significantly higher on it, relative to the participants of the other disciplines (p<0.05). There was no statistically significant difference between the scores of female and male participants, and across the various categories YoB. In terms of components, there was significant difference across the scores of the YoB categories for components 1, 11, 16, 18, and 19 (p<0.05). For components 1 and 11, year 1994 was significantly higher than the rest of the YoB categories; year 1995 higher for 16, year 2000 for 18, and year 1998 for 19.

In the post-assessment, the nursing participants scored significantly higher than those of the rest of the disciplines in the Roles and responsibilities subscale, only (p<0.05). Moreover, there was no statistically significant difference between the scores of female and male participants, and across the various YoB.

## Discussion

The current study revealed that the IPE intervention under investigation significantly increased the level of readiness to engage in cross-disciplinary learning and collaborations among the participating health professions’ students. This intervention constituted a novel experience for most of the participating students. Its efficaciousness in the context of the study encourages other universities in the region to adapt such interventions to increase students’ level of readiness to engage in cross-disciplinary learning and collaborations. Moreover, the evaluative tool utilized in this study, namely: RIPLS (21), proved to be internally reliable and externally valid in evaluating the students’ responsiveness to IPE.

There is increasing interest in the theoretical underpinning of IPE. Several key theories have been used in IPE curriculum design, many of which take a constructivist approach. SLT is one such example. Constructivism emphasizes how social encounters influence learners’ understanding and development (22). SLT emphasizes that learning occurs in the context of the experience, and places great emphasis on relationships and interactions with others in building understanding and developing the role of the individual within the greater community (23).

Similarly, RC argues that for a team to be effectively coordinated, there is a need for shared knowledge and understanding among its members, as well as for all the entailed relationships to be built upon shared goals and mutual respect (3). It is established in the literature that communication and other interpersonal skills among healthcare professionals are essential for the quality of healthcare delivery (24). Strong relationships in teams are expected to contribute to effective service delivery and improved patient health outcomes (25). In the setting of the current study, students of differing healthcare disciplines were able to collaborate and exchange knowledge. This enhanced their teamwork and fostered their professional identities, which is expected to translate, on the long run, in better outcomes of care.

Nursing students appear to significantly benefit from such IPE interventions. Relative to that of the rest of the disciplines, the level of readiness for inter-disciplinary teamwork among nursing students, in relation to the roles and responsibilities, increased the most after the intervention under investigation in this study. Along those lines, a previously conducted study revealed a significant improvement in RIPLS scores with nursing students following an interdisciplinary learning intervention but not for any of the other disciplines evaluated (26). For the IPE session in the current study, medical students were intentionally chosen in their pre-clinical years to try and improve the level of interaction between medical students and students of other health care disciplines. Non-medical students often find it difficult to achieve equal interaction as they expect medical students to lead the conversation due to their perception of them as ‘doctors’. This narrative is particularly prevalent in the Middle East and presents a greater incentive for breaking down the silos of different disciplines (27).

It is established that readiness for interprofessional learning is fundamental to healthcare team development (28). Hence, identifying factors that influence the students’ readiness for interprofessional learning is fundamental to developing learning strategies targeted to improve teamwork, quality of care, and ultimately patient health outcomes (26). An evaluation of attitudes towards IPE, among pharmacy students in the MENA region, showed that most respondents perceive IPE to be important (15). Another similar study looking at the perception of students training in medicine, nursing, and laboratory sciences in the Kingdom of Saudi Arabia (KSA) showed similar findings. It revealed that most respondents consider themselves to be ready for engaging in multidisciplinary collaborations (27).

The receptiveness to IPE tends to vary across disciplines and from one context to another. In this study, prior to the intervention, participants training in medicine scored higher in terms of readiness for cross-disciplinary work. This might be attributed to the longitudinal themes that are integral to most of the contemporary medical programs (including the MBBS program that the medical students participating in this study are enrolled in), where competencies such as professional identity and practice (which includes but is not limited to interprofessional practice) are systemically nurtured throughout the respective programs. Another study showed that pharmacy and dietetics students demonstrated a higher level of readiness for interdisciplinary learning compared to other disciplines (26). In a study conducted among South Korean health profession students, the nursing students’ perception of the importance, preference, and effectiveness of IPE was the highest, whereas medical students’ perception was the lowest. All students (i.e., medical, nursing, and pharmacy) perceived their present level to be lower than that required for each interprofessional competency (29).

As previously outlined, specific subscales of the RIPLS include teamwork and collaboration, professional identity, and roles and responsibility. Comparison of the subscale domains by discipline is useful in understanding views and attitudes that may be characteristic of a specific discipline and may provide an explanation for the differences observed between the disciplines for RIPLS composite score. The intervention under investigation significantly raised the level of readiness regarding teamwork and collaboration. It also increased the readiness in relation to acquisition and effectiveness of teamworking skills. Yet, there was no significant difference in relation to the need for positive relationship between professionals and other healthcare students. These findings are in alignment with the work of Lairamore et al. (2013) who reported improved RIPLS scores following a case-based interprofessional intervention across all disciplines (8).

It is well established that teams offer the promise to improve clinical care because they can aggregate, modify, combine, and apply a greater amount and variety of knowledge to make decisions, solve problems, generate ideas, and execute tasks more effectively and efficiently than any individual working alone (30). Accordingly, introducing students to interdisciplinary collaboration early-on in their careers will help foster the concept of a team mentality and comprehensive patient care, where each medical professional holds a piece to the puzzle.

Interventions such as the one investigated in this study holds the potential to significantly foster the participants’ professional identity. This appeared to be the case through decreasing their negativity around the subject matter and improving the extent of positivity associated with their identity due to such cross-disciplinary interventions. Interestingly, although the intervention improved the state of readiness among the participants in relation to the first two subscales: teamwork and collaboration, and professional identity, as mentioned above, there was no statistical significance around roles and responsibilities. The only exceptional component of this scale was: “The function of nurses and therapists is mainly to provide support for doctors”. Along those lines, another study aimed at evaluating the medical students’ readiness and perception of IPE in a medical college in the KSA revealed a similar pattern and recommended for shared academic events to focus on clarifying the roles and responsibilities of medical students in multi-disciplinary healthcare teams (27).

The current study is characterized by a few limitations, which can be perceived as opportunities for further research. This investigation was deductive in nature and relied solely on quantitative data. It would be useful for future studies to be exploratory in nature, where focus group sessions can be conducted to better understand why there are differences across healthcare disciplines in terms of receptiveness to IPE. Moreover, the study results are based on a single intervention that was conducted in two rounds for different batches. Although they were meant to be congruent, slight variations were inevitable. In addition, to maintain the participants’ anonymity, there were no personal identifiers recorded in the pre- and post-assessments of this study. This disabled linking the data, on the participant-level, which is why this study’s analysis was performed on an aggregate level. It is recommended for future studies to be based on a longitudinal design where the pre- and post-assessment are linked perhaps through assigning unique identifying numbers to the participants (which will maintain their anonymity). For the participants of the current study, the researchers are intending to conduct a follow-up investigation to assess how this IPE intervention affected their engagement in cross-disciplinary teamwork and collaborations in their clinical years and internships, and as they progress in their career paths. Also, the data collection was not electronic and relied on hardcopies of the respective surveys. Consequently, not all participants responded to all questions.

## Conclusion

IPE interventions, such as the one under investigation, hold the potential to significantly increase the participating health profession students’ level of readiness to engage in cross-disciplinary learning and collaborations. It is recommended for other universities in the MENA region to adapt IPE to improve future health professionals’ capacity to develop shared understanding and mutual respect within cross-disciplinary teams, which ultimately feeds into improved quality of care and patient outcome.

## Data Availability

All data is fully available without restriction

## Acknowledgement

The authors would like to extend gratitude to Professor Ayman Noreddin, Dr. Jaqueline Dias, and Dr. Ibrahim Mustafa for their support in recruiting, and in turn enrolling into the study, pharmacy, nursing, and physiotherapy students from the University of Sharjah.

## Notes

### Competing Interest Statement

The authors have declared no competing interest.

